# Development and validation of a multiplex real-time qPCR assay using GMP-grade reagents for leprosy diagnosis

**DOI:** 10.1101/2021.10.04.21264517

**Authors:** Fernanda Saloum de Neves Manta, Thiago Jacomasso, Rita de Cássia Pontello Rampazzo, Suelen Justo Maria Moreira, Najua Zahra, Marcelo Ribeiro-Alves, Marco Aurélio Krieger, Alexandre Dias Tavares Costa, Milton Ozório Moraes

## Abstract

Leprosy is a chronic dermato-neurological disease caused by *Mycobacterium leprae*, an obligate intracellular bacterium. Timely detection is a challenge in leprosy diagnosis, relying on clinical examination and trained health professionals. Furthermore, adequate care and transmission control depend on early and reliable pathogen detection. Here, we describe a qPCR test for routine diagnosis of leprosy-suspected patients. The reaction simultaneously amplifies two specific *Mycobacterium leprae* targets (16S rRNA and RLEP), and the human 18S rRNA gene as internal control. The limit of detection was estimated to be 2.29 copies of *M. leprae* genome. Analytical specificity was evaluated using a panel of 20 other skin pathogenic microorganisms and *Mycobacteria*, showing no cross-reactivity. Intra- and inter-operator C_p_ variation was evaluated using dilution curves of *M. leprae* DNA or a synthetic gene, and no significant difference was observed between three operators in two different laboratories. The multiplex assay was evaluated using 97 patient samples with clinical and histopathological leprosy confirmation, displaying high diagnostic sensitivity (91%) and specificity (100%). Validation tests in an independent panel of 50 samples confirmed sensitivity and specificity of 97% and 98%, respectively. Importantly, assay performance remained stable for at least five months. Our results show that the newly developed multiplex qPCR effectively and specifically detects *M. leprae* DNA in skin samples, contributing to an efficient diagnosis that expedites the appropriate treatment.

**Author Summary:** Leprosy is a chronic dermato-neurological disease caused by *Mycobacterium leprae*, an obligate intracellular bacterium. Disease diagnosis is currently performed on skin examinations for clinical signs, bacilli staining in skin smears and invasive skin biopsies. However, the spectrum of clinical manifestations and the low bacterial load can hinder accurate diagnosis, which is critical for providing proper intervention and adequate care as well as for establishing transmission control. Quantitative PCR (qPCR) methods for detecting bacterial DNA are more sensitive and could aid in differentially diagnosing leprosy from other dermatological conditions. In this work, we present a new multiplex qPCR that detects two bacterial genes for the diagnosis and a human gene as an internal reaction control. The new qPCR, developed using GMP-grade reagents, is highly sensitive, specific, reproducible, and stable. The results presented here are the basis of a novel and robust tool with potential to increase the accuracy of leprosy diagnosis in routine or reference laboratories.

## Introduction

Leprosy is a neglected infectious disease that still represents a public health issue (1) with more the 200,000 cases every year worldwide. Diagnosis is generally late and, although a specific and effective treatment is available, it is likely that transmission occurs before the patient is diagnosed and adequately treated, thus contributing to sustained transmission. The high number of young patients (under 15 years old) and patients with disabilities due to advanced stage of the disease, confirms this hypothesis (1). Furthermore, clinical forms vary to a great extent, from localized (tuberculoid) to disseminated (lepromatous) forms, making diagnosis difficult. Evidence suggests that early diagnosis could prevent transmission and help epidemiological control (2).

Methods such as bacterial load detection by microscopy and histopathological examination have been the main complementary tools for the diagnosis of leprosy (2–4). Classical bacteriological methods cannot confirm leprosy since *M. leprae* does not grow *in vitro*. In addition, there is no reliable marker to estimate the risk of disease progression (5,6). In this regard, the sequencing of *M. leprae* genome (7) was a milestone towards the improvement of direct *M. leprae* detection, leading not only to better characterization of genomic targets unique to *M. leprae* strains but also to an extensive comparison of different mycobacteria.

At the time of the first sequences became available, the polymerase chain reaction (PCR) technique was laborious and very expensive, averting its universal application. However, as PCR was further developed, it became more affordable, versatile and reliable, with fully automated systems becoming commercially available from different companies (8–10). For tuberculosis, routine tests using PCR are reducing the turnaround time, allowing same day treatment initialization, which might impact resistance prevalence (11–13). Cost-effective nucleic acid detection assays are relatively widespread, but assays for some neglected diseases are still missing. In leprosy, the situation is even more difficult due to reduced and late investments directed to diagnostic tests (14).

In the last few years, many studies have been carried out using the PCR technique to detect *M. leprae* DNA in clinical specimens. PCR have been used especially under challenging diagnoses such as equivocal paucibacillary (4,15–18) or monitoring household contacts (19,20). In this context, several different targets have been described in an attempt to establish the most sensitive and specific assay (16,20–28). However, most of the PCR protocols were developed, evaluated, and validated using reagents or tests produced without good manufacturing practices (GMP). Also, most of the studies enroll only leprosy patients and do not recruit patients with other common dermatological diseases that are differential diagnosis to leprosy. Thus, the development and validation of an assay over different laboratories has become a necessity.

Here, we present the development and validation of a multiplex real-time PCR assay aiming to standardize the leprosy molecular diagnostic assay. The protocol was designed to simultaneously detect two *M. leprae* targets (16SrRNA and RLEP genes), previously used in several studies (4,16,19,26,29), and one mammalian target (18S rRNA gene), that serves as reaction control (30). Cross-reactivity was evaluated using DNA from 20 related mycobacterial and other skin pathogenic species, and no match was found. The new assay was validated using 97 skin biopsies and an independent panel enrolling 50 samples retrieved from patients previously characterized by clinical examination and histopathology, showed high sensitivity and specificity. The new multiplex PCR was also assessed for quality control standards and the data indicate that the assay is stable and reproducible. The results presented here are the basis of a novel and robust tool with potential to increase the accuracy of leprosy diagnosis in routine or reference laboratories.

## Material and methods

### Ethics statement

The Ethics Committee of the Oswaldo Cruz Foundation approved this study (CAAE: 38053314.2.0000.5248, number: 976.330–10/03/2015). Written informed consent was obtained from all patients 18 years or older, or from the parents/guardian of patients under 18.

### Clinical samples

Leprosy patients were enrolled from the Leprosy clinic from the Oswaldo Cruz Foundation in the city of Rio de Janeiro, Brazil. Skin biopsies were collected using a 6-mm punch and stored in 70% ethanol at -20 °C until processing.

Ninety-seven samples (53 skin biopsies from leprosy patients and 44 skin biopsies from patients with other skin diseases) were used for qPCR tests. Clinical and demographic characteristics of all patients are shown in Table 1.

**Table 1:**
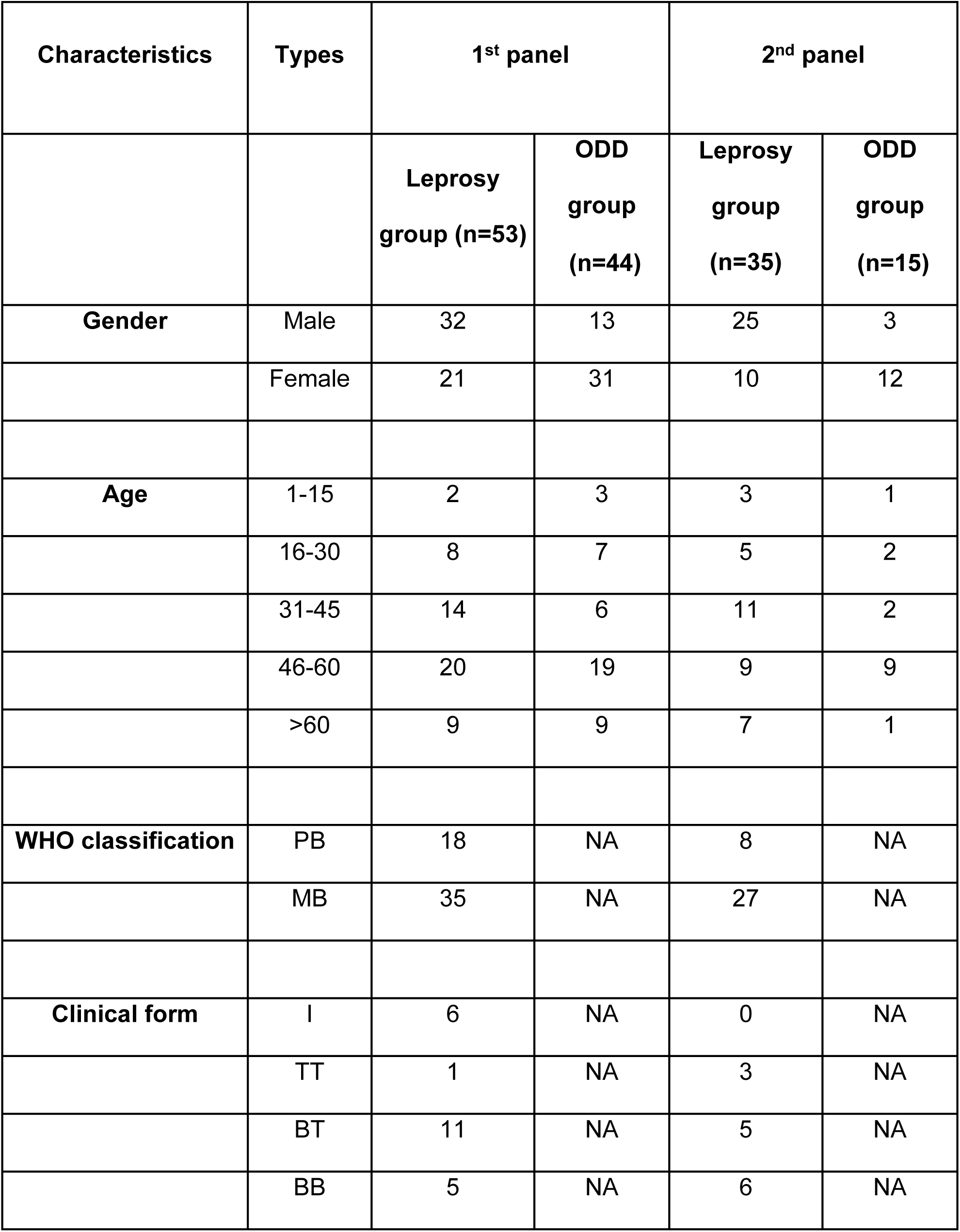
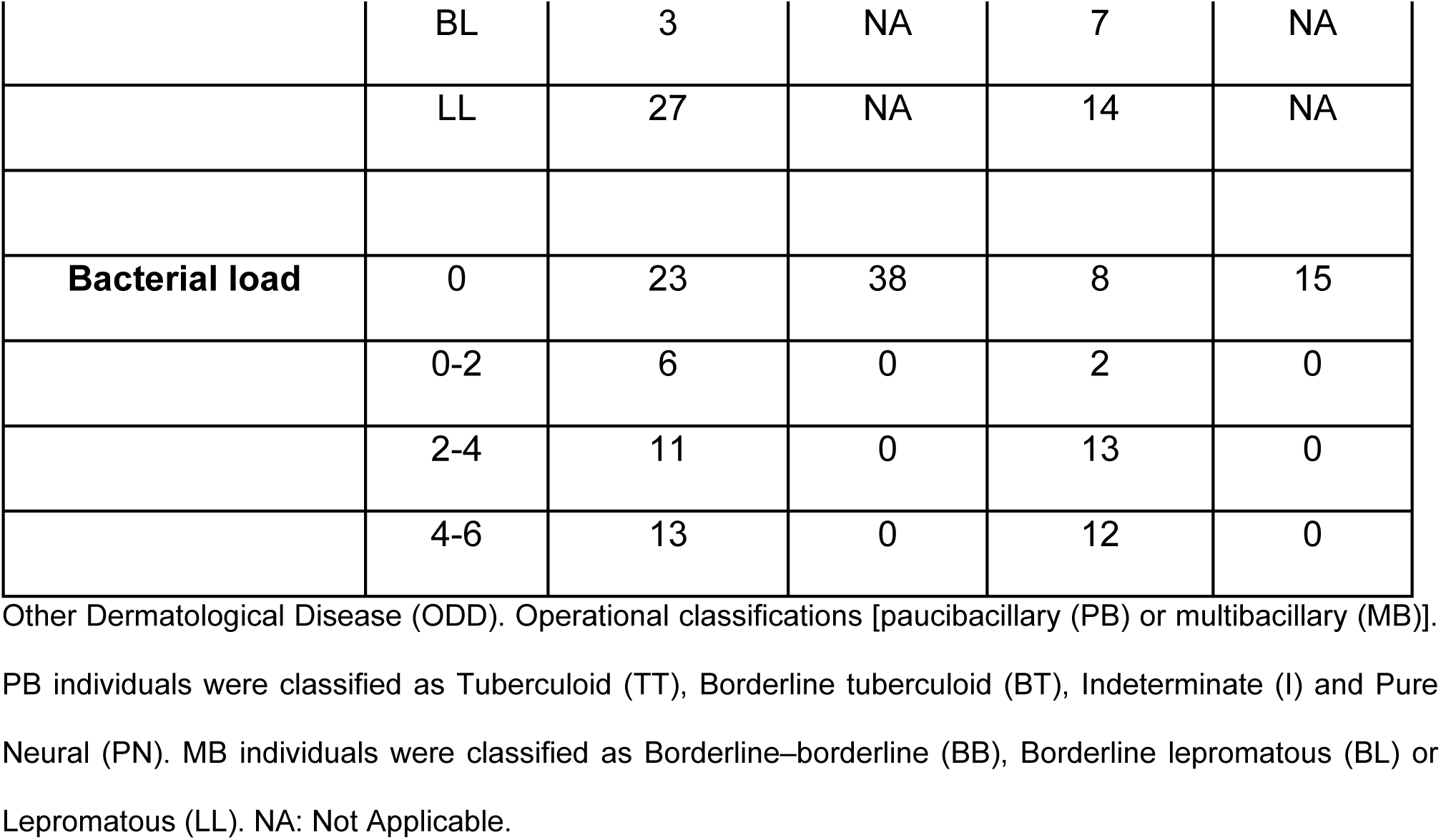
Clinical and demographic characteristics of the leprosy and other dermatological disease cases

Leprosy patients were defined according to the clinical, bacteriological, and histopathological Ridley-Jopling (R&J) classification and the operational classification in multibacillary (MB) or paucibacillary (PB) forms according to the WHO (31). Leprosy or other dermatological diseases (ODD) patients were treated according to their respective condition. Leprosy paucibacillary (PB) or multibacillary (MB) patients were treated according to the Ministry of Health recommendations, while ODD patients were treated accordingly for each specific disease.

### Replication Study

To validate the conditions and analysis parameters established with the clinical samples from Oswaldo Cruz’s Leprosy Clinic, we tested a distinct collection of 50 skin biopsy samples that were also obtained by the Leprosy Clinic. The second set of samples was sent to the Global Health Institute, École Polytechnique Fédérale de Lausanne, Switerzland, where DNA samples was extracted and was blindly characterized by conventional PCR according to a previously published protocol (32). The extracted DNA was then sent back to the Leprosy Clinic at Oswaldo Cruz Foundation, where it was blindly analyzed with the qPCR developed in the present study. After both PCR analyses were performed, blinding was removed and the results were compared. Of these 50 samples, fifteen samples were from patients with other skin diseases, 27 patients MB leprosy and eight from PB leprosy. The group presented a 1.27:1 ratio of males to females. The mean age was 44.8 (+/− 17.72 SD), and the range was 8-77. Details on the clinical characteristics are shown in supplementary table 1.

### Mycobacterial isolates samples

*M. leprae* Thai-53 purified from athymic BALB/c (*nu/nu)* mouse footpads was kindly provided by Dr. Patricia Rosa at the Lauro de Souza Lima Institute, Bauru, São Paulo, Brazil. Purified DNA from *M. leprae* was used as positive control and in analytical sensitivity studies.

DNA from 21 mycobacterial samples were used for the analytical specificity study. *L. amazonensis* and *L. braziliensis* was kindly provided by Dr Elisa Cupolillo by the Laboratório de Pesquisa em Leishmaniose (IOC-Fiocruz) and *M. avium, M. gordonae, M. manteni, M. africanum* subtype I, *M. africanum* subtype II, *M. bovis, M. bovis* (BCG), *M. canettii, M. fortuitum, M. gordonae, M. intracellulare, M. kansasii, M. microti, M. pinnipedii, M. simiae, and M. tuberculosis* were kindly provided by Dr. Phillip Suffys at Laboratório de Biologia Molecular Aplicada a Micobactérias (IOC-Fiocruz).

### Synthetic DNA

The synthetic DNA (gBlock^®^) was purchased from Integrated DNA Technologies (IDT) and consists of a double-stranded DNA containing the sequences of the three genomic targets (RLEP, 16S, and 18S) (S1 Appendix). The lyophilized DNA was reconstituted to 10 ng/μL (corresponding to 1.83 × 10^9^ copies per reaction) in TE pH 8.0, following the supplier’s protocol.

### DNA extraction

DNA extraction from the biopsies was carried out using DNeasy Blood and Tissue^®^ extraction kit (Qiagen, Germany). The total extracted DNA was quantified with NanoDrop^®^ (Thermo-Fisher Scientific, Waltham, MA, USA) and stored at -20 °C. *M. leprae* DNA from nude mice footpad was purified using TRIzol reagent (Life Technologies, Carlsbad, California) following the manufacturers’ instructions, as previously described (3). DNA used in the replication study were extracted using QIAmp UCP Pathogen Mini kit (Qiagen GmbH, Hilden, Germany).

### Standard curve and 95% limit of detection (LoD_95%_) assessment

The standard curve was used for determination of the limit of detection and assay stability. A series of 10-fold dilutions was prepared from either *M. leprae* or synthetic DNA, using DNA purified from human blood obtained from healthy donors as matrix. The dilution series used for the standard curve and the LoD_95%_ determination spans concentrations from 500 ag/reaction to 5 ng/reaction of purified *M. leprae* DNA, and 1.83 to 1.83 × 10^7^ copies/reaction (equivalent to 0.5 ag/reaction and 5 pg/reaction, respectively) of synthetic DNA.

### Quantitative PCR (real-time PCR assays)

A multiplex real-time qPCR assay targeting simultaneously two *M. leprae* regions and an internal reference human sequence was developed. The primers and hydrolysis probes were designed to detect regions from RLEP and 16S rRNA genes (29) from *M. leprae*, and the human 18S rRNA (30) (Table 2). Reactions were performed on an ABI7500 Standard instrument (Thermo-Fisher Scientific, Waltham, MA, USA), using Multiplex PCR Mastermix (IBMP/Fiocruz PR, Curitiba, Brazil). For each reaction, 5 μL of DNA solution was added for a 25 μL final volume. Reaction mixtures were prepared in triplicates and amplified at 95 °C for 10 min, and 45 cycles of 95 °C for 15 sec and 60 °C for 1 min. All reactions included a positive control (mouse foot-pad *M. leprae* DNA and/or high-bacterial load lepromatous leprosy patient purified DNA), and water as a non-template control (NTC; PCR reaction without any template DNA).

**Table 2:**
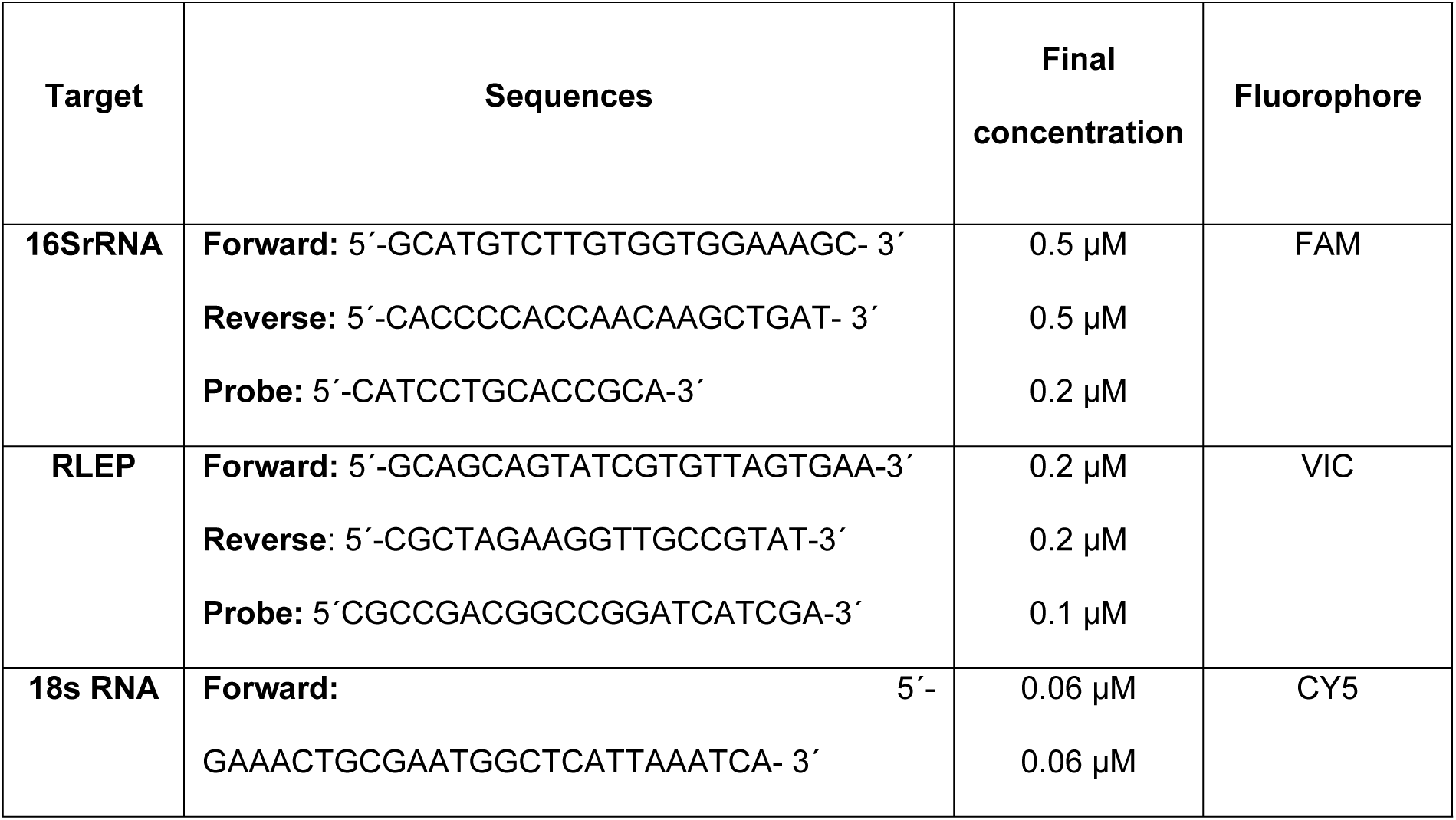
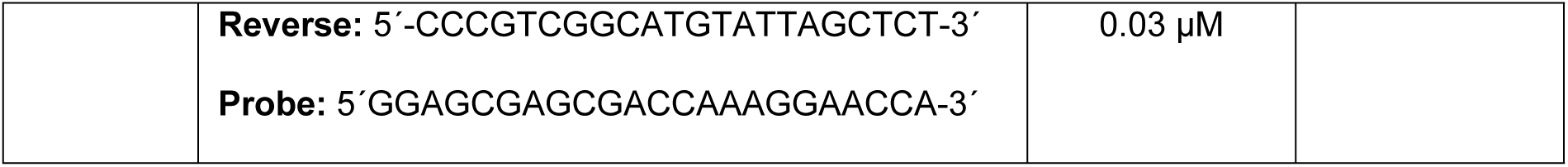
Sequences, concentration, and fluorophores of the oligonucleotides contained in the multiplex qPCR assay.

### Stability

The stability of the new multiplex qPCR was evaluated the synthetic DNA template diluted in TE to the concentrations of approximately 2 × 10^8^, 2 × 10^7^, 2 × 10^6^, 2 × 10^6^, 2 × 10^5^, 2 × 10^4^, 2 × 10^3^, 2 × 10^2^, 2 × 10^1^, 10, 5, and 2.5 copies per reaction.

All reagents (oligomix 25X and qPCR mix) were maintained in independent aliquots at -20 °C at the Leprosy Laboratory (Fiocruz-RJ). Tests with the dilution series described above were repeated weekly for the first month, and then once a month for five months.

### Data Analyses and Statistics

Qualitative (diagnostic sensitivity and specificity, accuracy) and quantitative (intra- and inter-laboratory repeatability and reproducibility, analytical sensitivity and specificity) validation tests were performed. The 95% limit of detection (LoD_95%_) was calculated by fitting a Probit model to the estimated detection probabilities. Data were processed and analyzed using customized scripts for R version 3.5.1 (downloaded from http://www.Rproject.org/).

## Results

### Analytical performance

Primers and hydrolysis probes designed to target 16S rRNA and RLEP sequences of *M. leprae* were tested in multiplexed reactions to concomitantly detect the human 18S rRNA sequence. Optimal fluorescence thresholds were chosen based on the common practice that it should be positioned on the lower half of the fluorescence accumulation curves plot from the 10-fold dilutions, crossing most if not all fluorescence signals on the exponential segment of the curve on a logarithmic scale (figure 1). Therefore, after setting the baseline to the automatic function, fluorescence threshold values chosen for determining C_p_ (Crossing point) values for each target were set to intercept the positive controls and avoid the negative ones, being established as follows: 0.2 for RLEP, 0.15 for 16S rRNA, and 0.16 for 18S rRNA.

**Fig 1.**
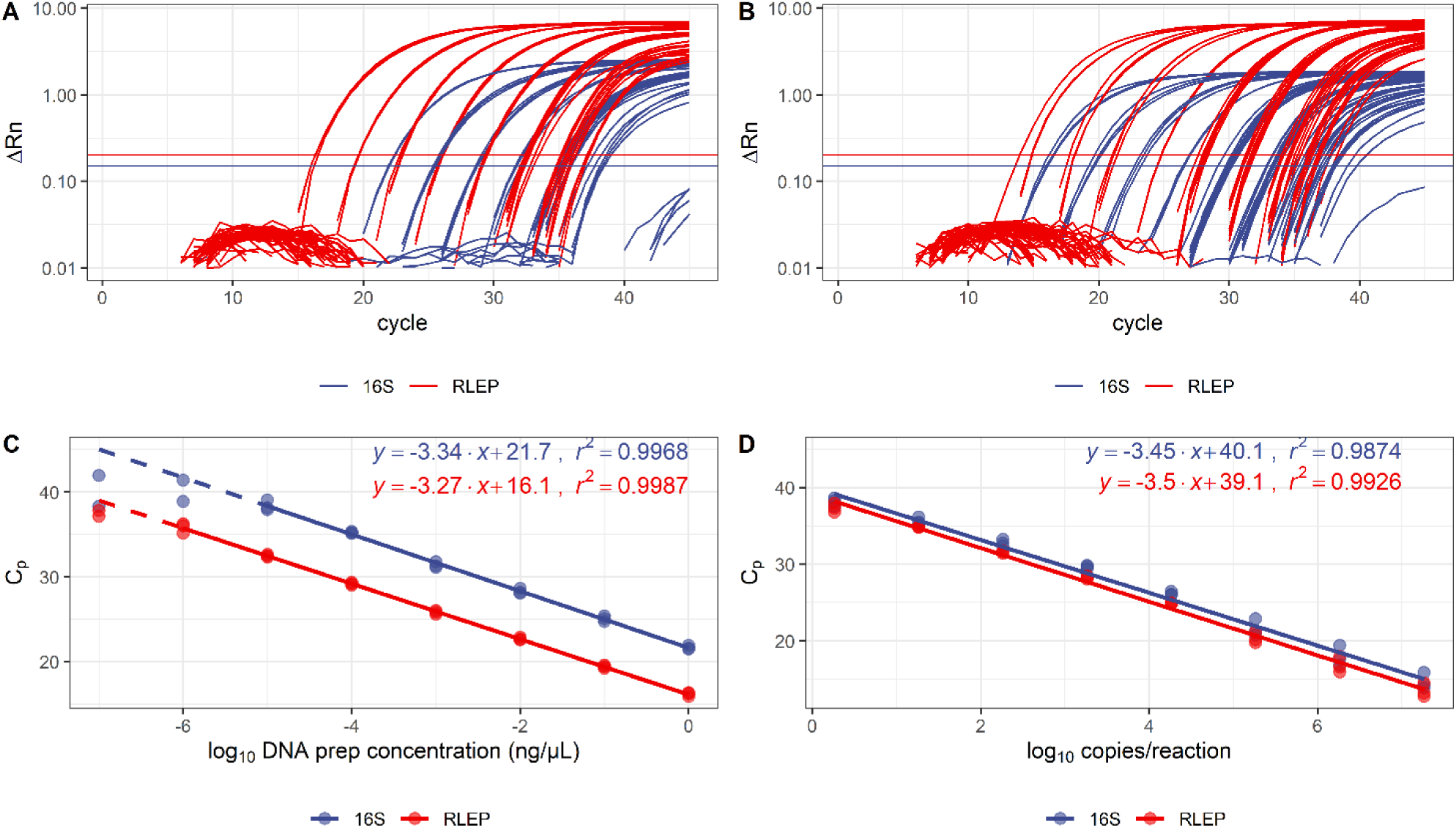
Standard curves of the amplification of 16SrRNA and RLEP targets in *M. leprae* DNA and in a synthetic construct. Panels A and C show the calibration curves obtained using *M. leprae* DNA, diluted in total DNA extracted from *M. leprae*-negative whole blood. Continuous lines show the linear range and the dashed lines are extrapolations towards the non-linear range. Efficiencies calculated from the linear ranges were 99.2% for 16SrRNA and 102.2% for RLEP, and r^2^ were 0.9968 and 0.9987, respectively. Panels B and D show the calibration curves obtained using a synthetic gene containing one copy of each target per molecule, diluted in total DNA extracted from *M. leprae*-negative whole blood. The efficiencies were 94.9% for 16SrRNA and 93% for RLEP, and r^2^ were 0.9874 and 0.9926, respectively.

The analytical 95% limit of detection (LoD_95%_) was determined from a series of tests in which DNA extracted from *M. leprae* was diluted from 5 ng to 100 ag/reaction. Figure 2 shows the fitted Probit models and the obtained LoD_95%_ for 16S rRNA and RLEP, which were experimentally determined as approximately 450 fg of DNA (ca. 126 *M. leprae* genomes) for the 16SrRNA gene and about 4.60 fg of DNA (ca. 1.3 *M. leprae* genomes) for the RLEP gene.

**Fig 2.**
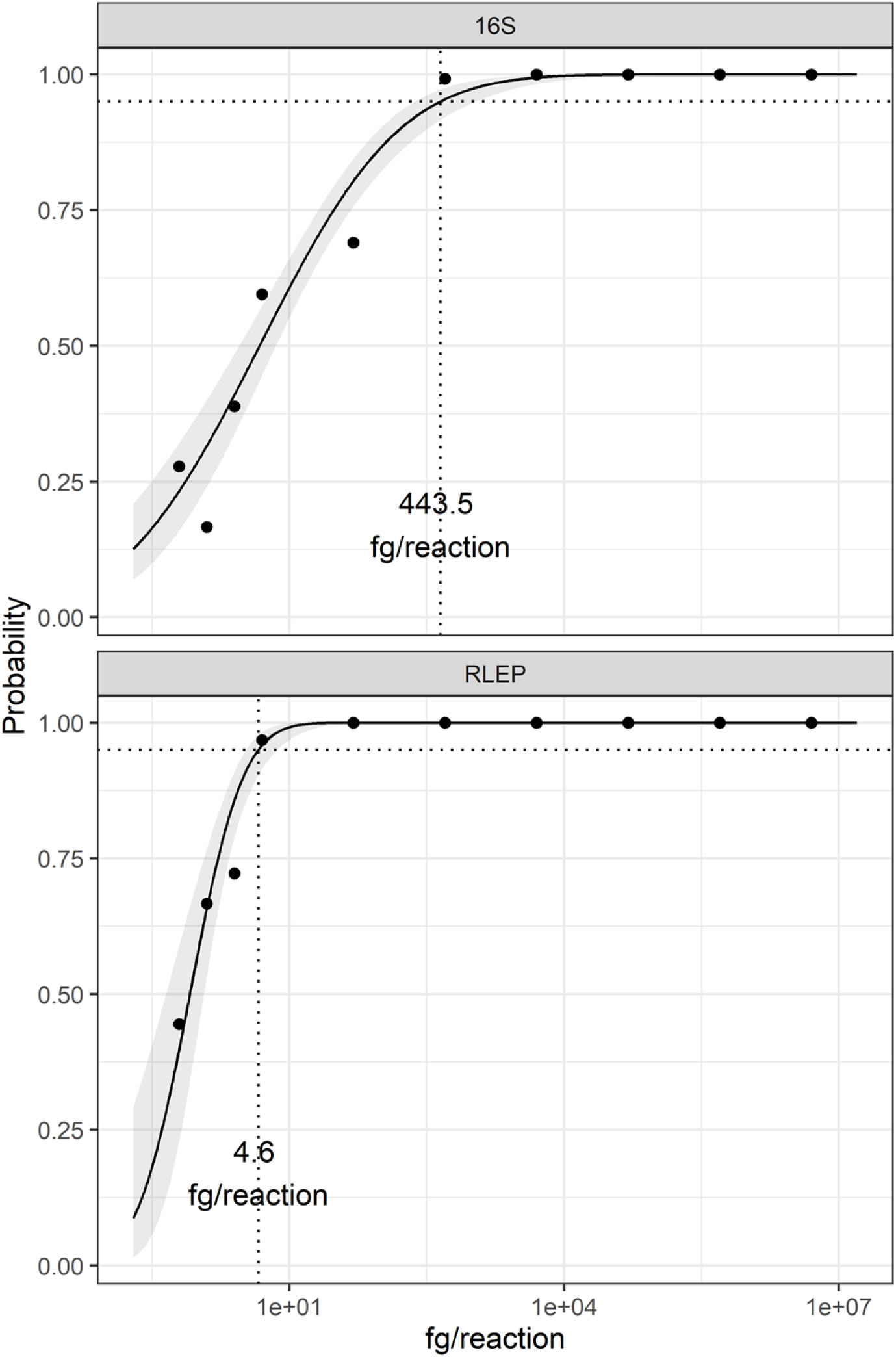
Analytical 95% limit of detection (LoD_95%_) for 16SrRNA and RLEP in multiplexed qPCR. *Mycobacterium leprae* DNA was diluted in DNA extracted from whole blood from healthy donors and tested from 5 ng to 0.5 fg/reaction. Probability of detection was calculated for 16S and RLEP (tom and bottom panels, respectively) from nine independent experiments, and a Probit model was fit to the data (black lines). The grey ribbon around the model fit indicates the 95% CI on the predicted probability. Dotted lines indicate the interpolation to determine the concentration at a 95% probability. The calculated LoD_95%_ is displayed on each plot in femtograms of DNA/reaction.

The developed multiplex reaction was evaluated against a collection of microorganisms to assess the specificity of the primers and probes under these conditions. The selection included several mycobacteria, as well as a few other pathogens associated with skin diseases such *Leishmania* (figure 3). We only considered any species as cross-reactive if all the technical replicates displayed amplification for at least one of the targets which was not the case for any of the species tested. Most positive amplifications observed correspond to RLEP, which was detected in two out of three replicates in *M. fortuitum* and *M. kyroniense*. Even though some reactions presented 16S rRNA signals above the threshold, these amplifications are very uncharacteristic and are easily distinguishable from a proper amplification when compared with the positive control with 500 fg/reaction of *M. leprae*.

**Fig 3.**
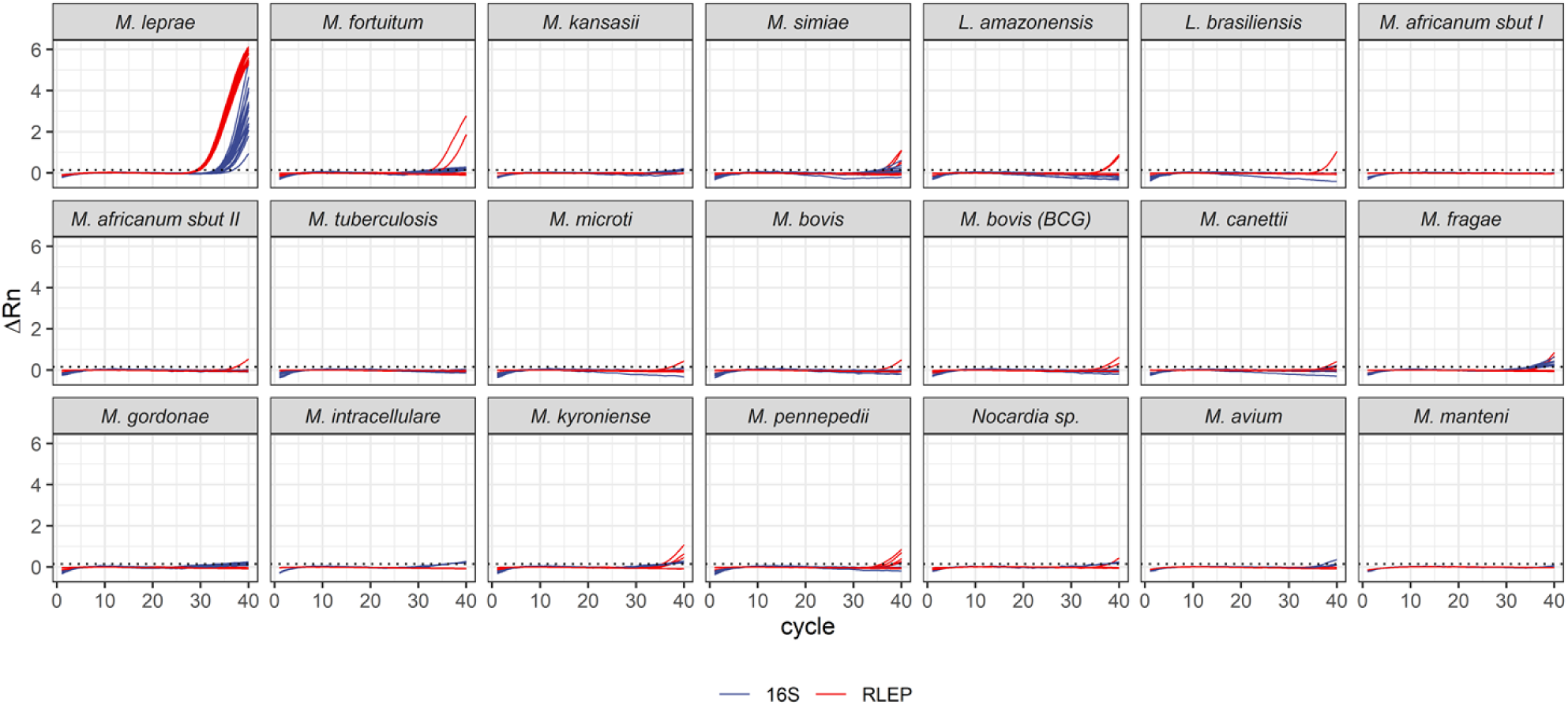
Analytical specificity for the 16SrRNA and RLEP multiplexed reactions. Extracted DNA from the indicated microorganisms (5 ng/µL each) were used in the multiplexed reactions performed in technical triplicates in two independent experiments. Results are compared to the amplification plot for 100 fg *M. leprae* DNA/µL (top-left panel). Amplification profiles are shown for each target, and each line corresponds to one individual well. The dotted lines indicate the threshold for RLEP (which is the highest of the two *M. leprae* targets, at 0.2).

### Repeatability and reproducibility

Three independent operators performed three replicate runs each, in consecutive days, and evaluated the repeatability and reproducibility of the multiplex reactions. For each replicate, a new dilution series for the synthetic gene was prepared from a concentrated aliquot to be used as template. Table 3 shows the relative deviations observed within and between operators, respectively. The data shows that all intra-operator replicates were remarkably reproducible, with only one point (Op. 1, 16S, 1.83 × 10^2^) displaying a relative standard deviation (rRSD%) above 5%, but still well below 10%. The inter-operator variability was also very low, and the largest variation was observed for the 16S target. Nonetheless, the rRSD% was between 1.38 and 11.57 across the dilution range, which shows an excellent reproducibility for a quantitative test (see also supplementary table 2).

**Table 3.**
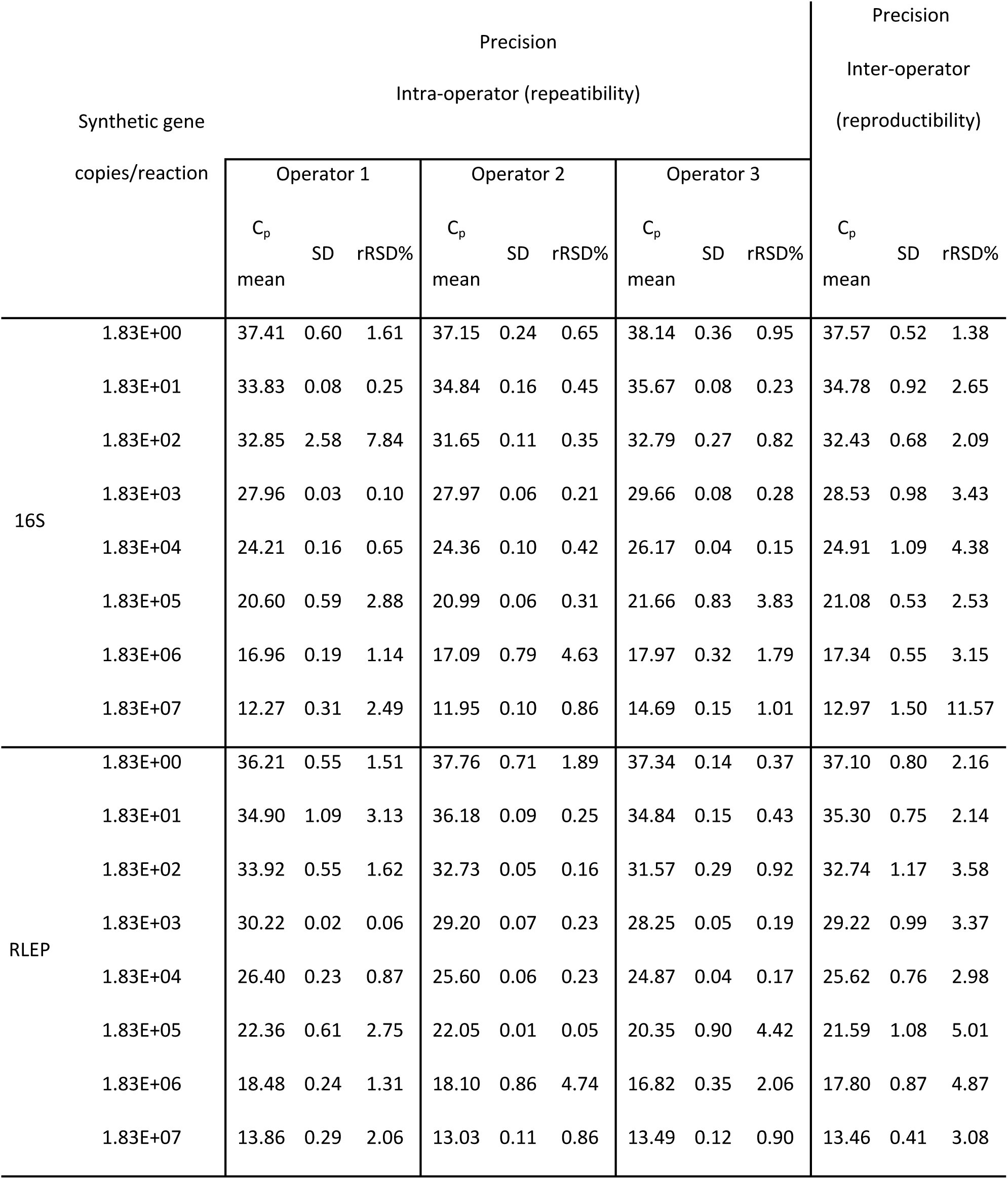
Precision measurement for repeatibility and reproductibility.

The accuracy of the determinations performed by the multiplex real-time qPCR assay was also estimated using the synthetic DNA. To evaluate the intra- and inter-repeatability (or intermediate precision) for operators, we calculated the arithmetic mean, standard deviation, and relative standard deviation percentage of three independent experiments. It is noteworthy that the detection of the human target 18S rRNA does follow the same dilution trend for the other targets because the synthetic template was not diluted in human DNA.

In summary, for both *M. leprae* targets we observed that all points showed excellent reproducibility and repeatability. As expected, detection of the human target 18SrRNA loses reproducibility as it becomes scarce in the reaction due to the dilution factor. It is noteworthy that there is no variation in the detection of the human target 18S rRNA when *M. leprae* DNA was present in the synthetic control molecule, i.e., in a 1:1 ratio, supporting the notion that the multiplexed reactions do not interfere with each other (data not shown).

### Stability

Storage stability was assessed by performing monthly evaluations of reactions with different concentrations of the synthetic DNA molecule for 5 months. Most of the data points tested varied below the established limit of three standard deviations above the average of all time points. Figure 4 shows the C_p_ obtained for the three evaluated targets (16SrRNA, RLEP, and 18SrRNA) in representative concentrations for brevity, over a 5-month period. The test remained reliable for the entire range of concentrations tested.

**Fig 4.**
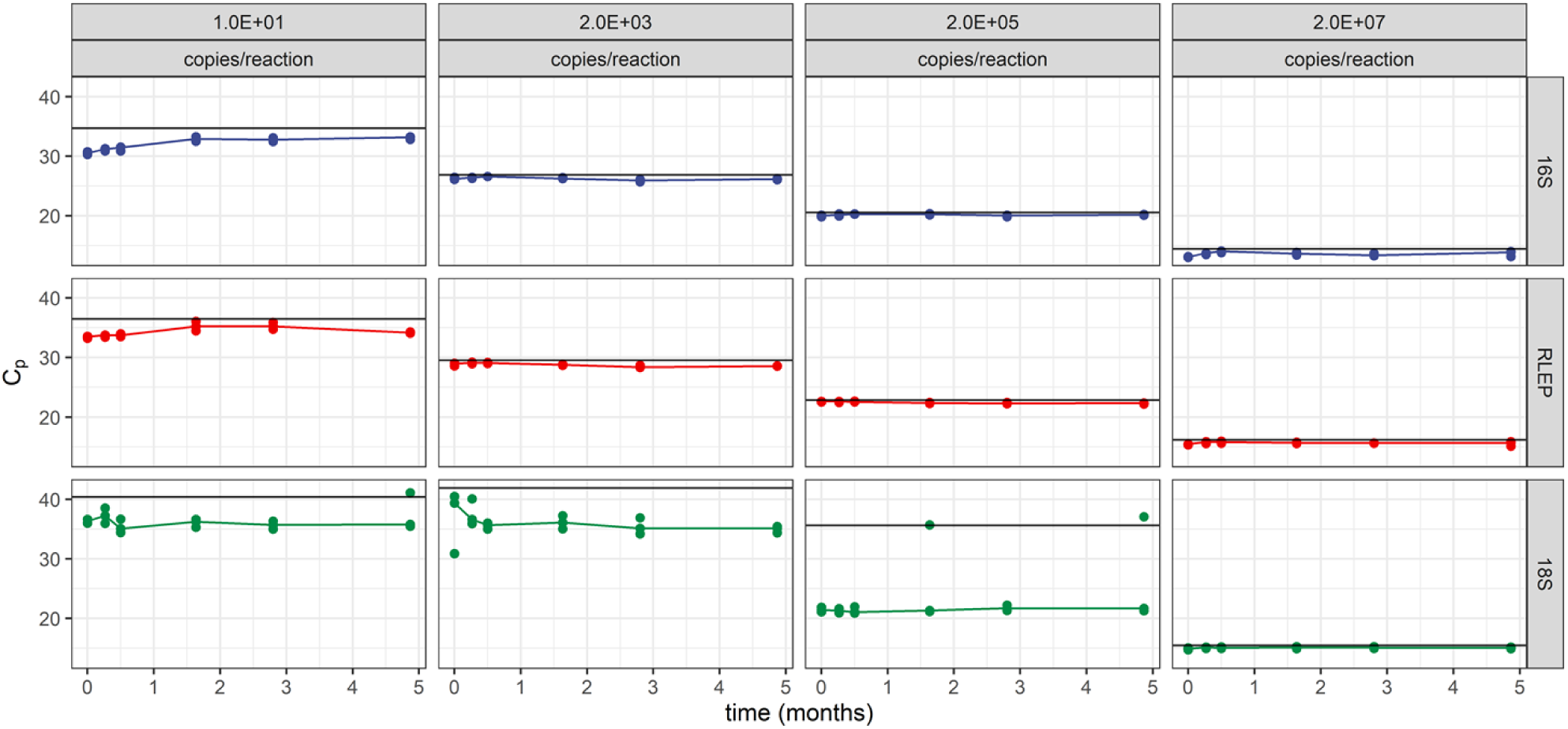
Stability of the reactions over five months using synthetic DNA as a template. Each panel shows the C_p_ values obtained for each target (lines of panels) and for each template concentration (columns of panels) over time. Points represent one technical replicate. Black horizontal lines indicate the upper tolerance limit defined as three standard deviations above the mean C_p_ for each template concentration.

### Diagnostic performance

The implemented setup involved the interrogation of two target sequences from *M. leprae* to classify clinical samples correctly while mitigating possible false positives. To evaluate the diagnostic performance, we first established optimal parameters for the analysis, considering possible cross-reactions that may occur in the laboratory routine. Figure 5 shows the receiver operating characteristic (ROC) curves for a subset of the best-performing combinations of cutoff values for 16SrRNA and RLEP. Data for the full range of C_p_ cutoffs combinations are listed in the supplementary table 3.

**Fig 5.**
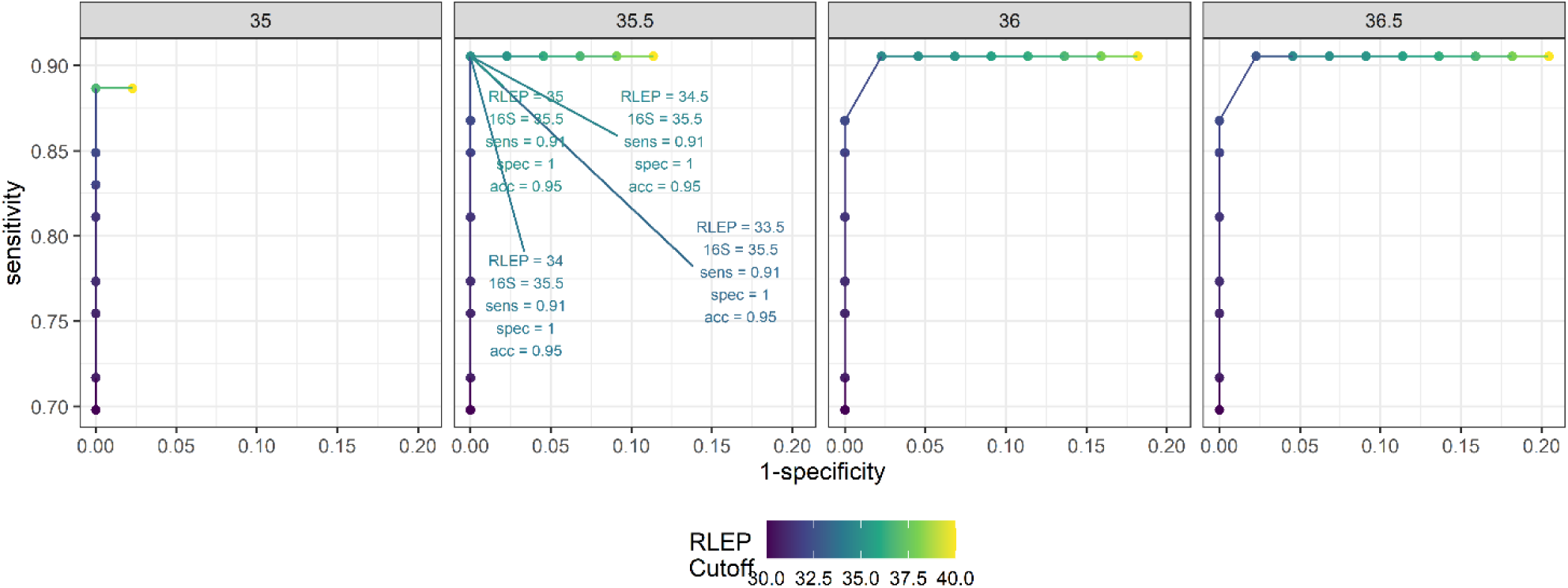
Diagnostic performance of the new multiplex qPCR. Different combinations of cutoff values for 16SrRNA (panels) and RLEP (color scale) were tested on a patient panel (n = 97). For each combination of cutoff values, the sensitivity and specificity were calculated and plotted as ROC curves. Here, only C_p_ cutoff values for 16SrRNA between 35 and 36.5 are shown. The combinations resulting in a specificity of 1 and the highest sensitivity for each condition are annotated.

Based on these results, the best combination of cutoff values (35.5 for 16SrRNA and 34.5 for RLEP) showed a sensitivity of 91% and specificity of 100%. These parameters were used to establish the decision algorithm presented in Table 4.

**Table 4.**
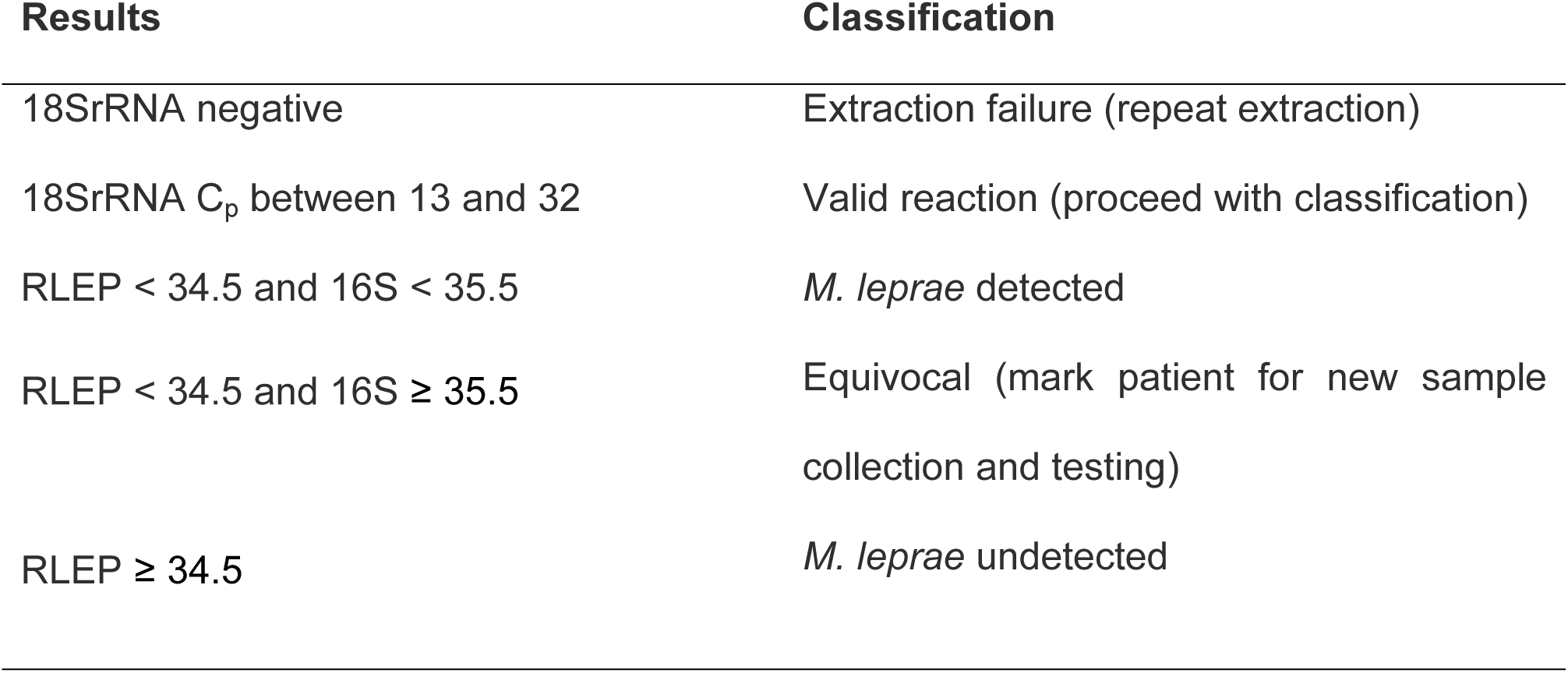
Decision algorithm for classification of samples based on the data obtained with the new multiplex qPCR.

| Results | Classification |
| --- | --- |
| 18SrRNA negative | Extraction failure (repeat extraction) |
| 18SrRNA C<sub>p</sub> between 13 and 32 | Valid reaction (proceed with classification) |
| RLEP < 34.5 and 16S < 35.5 | *M. leprae* detected |
| RLEP < 34.5 and 16S ≥ 35.5 | Equivocal (mark patient for new sample collection and testing) |
| RLEP ≥ 34.5 | *M. leprae* undetected |

Next, the molecular diagnosis obtained using the new multiplex PCR, was compared to the clinical diagnosis of each sample (Figure 6 and supplementary table 4). Results show that the qPCR reaction and classification algorithm correctly characterized 48 of the 53 samples previously described as “Leprosy” by the clinical outcome. Of the 5 misclassified samples, one was classified as negative for *M. leprae* and four were in the “equivocal” quadrant. The Bacterial load for the 5 misclassified samples were 0.

**Fig 6.**
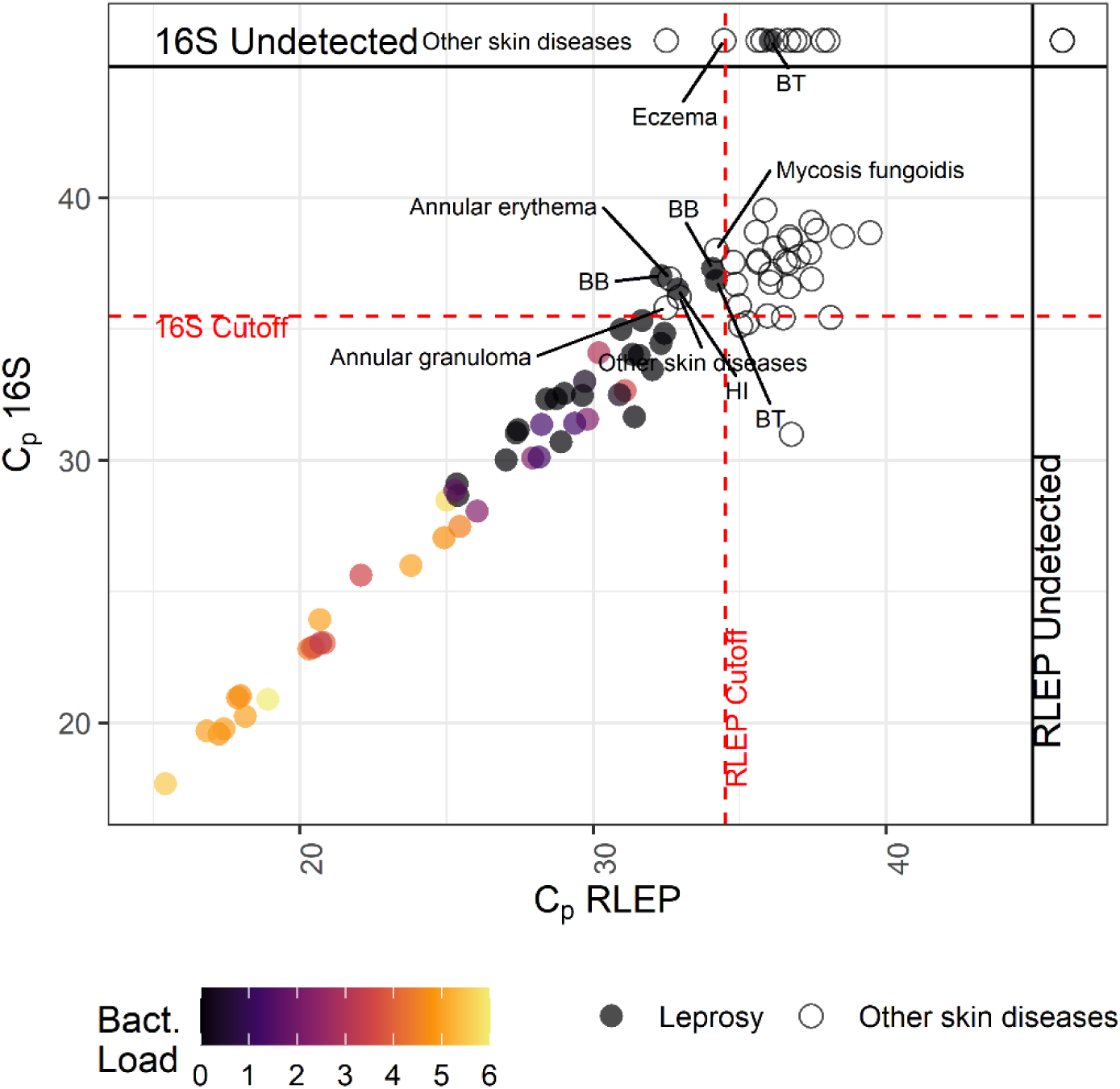
Distribution of C_p_ values obtained for the training panel. Each point represents a different sample (mean C_p_s of a technical duplicate). Filled circles represent leprosy samples and open dots represent negative samples, as defined by the clinical assessment. Points aligned to the top and right margins indicate samples in which 16SrRNA or RLEP, respectively, were not detected within 45 cycles. Bacterial load is show as a color gradient (samples for which bacterial load information was not available are filled in grey). Dotted red lines indicate the cutoff values from Table 3. Equivocal or misclassified samples are annotated with the operational classification (false negatives) or with the diagnosis for clinic-negative samples.

None of the 44 samples characterized as “Other skin diseases” were classified as *M. leprae*-positive by our reaction and decision algorithm. Thirty-eight of these samples were classified as “Negative” and 6 as “equivocal”.

### Assay validation

Conditions established with the training cohort were tested on an independent set of samples, which were previously characterized using a distinct qPCR method described in Girma et al. (32). The comparison between the original classification and the new results is shown in figure 7 and supplementary table 1. We tested 50 samples, of which 34 were previously characterized as positive and 16 as negative.

**Fig 7.**
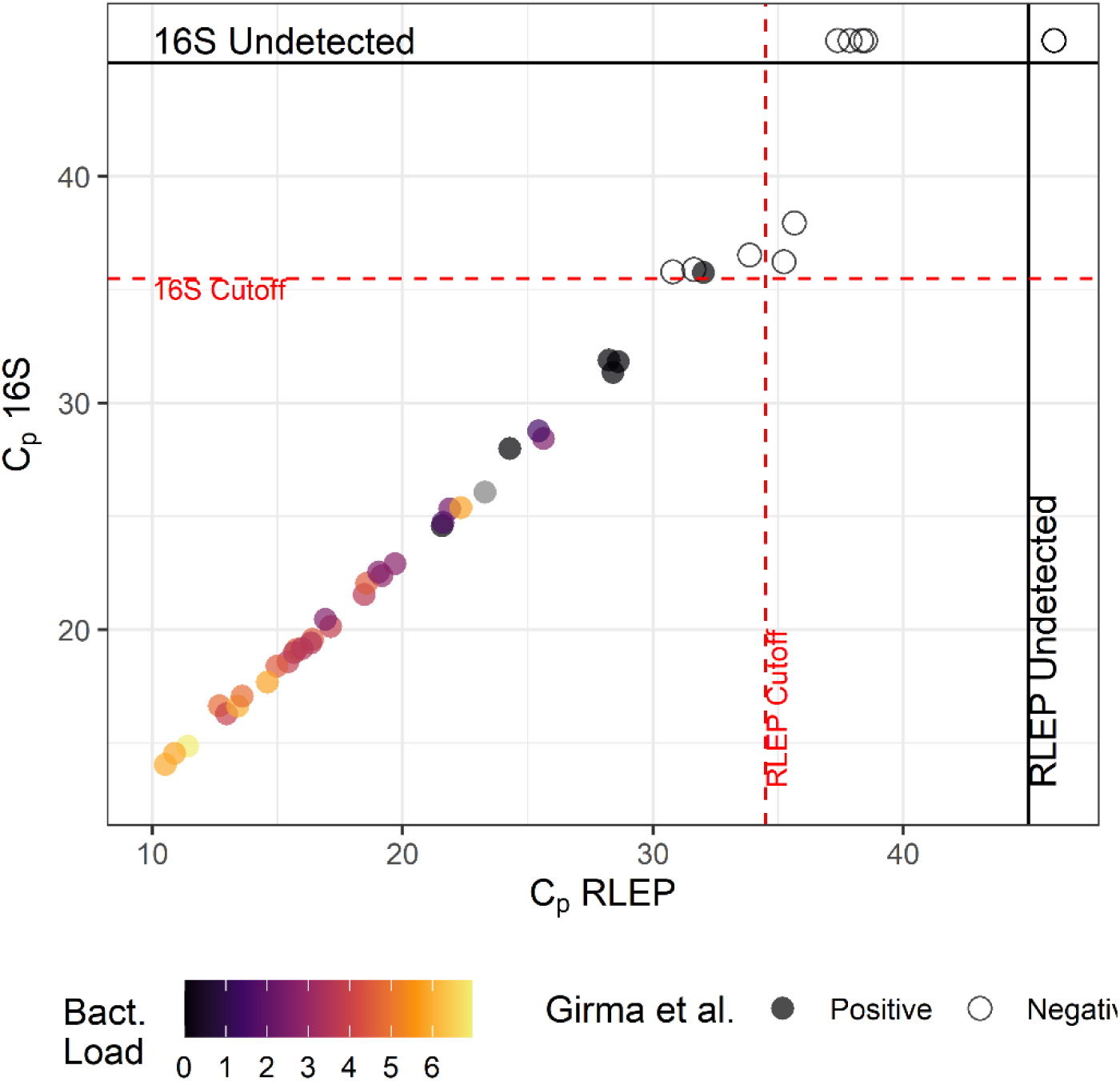
Validation of parameters on a in an independent panel. **P**reviously characterized validation samples were subjected to the new qPCR described in the present study. Each point represents a different sample. Filled circles represent leprosy samples and open dots represent negative samples, as defined by Girma *et al*. Points aligned to the top and right margins indicate samples in which 16SrRNA or RLEP, respectively, were not detected within 45 cycles. Bacterial load is show as a color gradient (samples for which bacterial load information was not available are filled in grey). Dotted red lines indicate the cutoff values from Table 3.

The 50 samples were classified according to our algorithm, resulting in 33 correctly classified as positive and 11 correctly classified as negative. Of the four samples classified as equivocal, two were negative for the reference method and one was positive according to Girma et al (32). The sensitivity, specificity and accuracy calculated for this sample set were 97.1%, 100% and 98%, respectively.

## Discussion

Leprosy is a chronic infectious disease presenting great diversity of clinical forms with distinct immunological and histopathological features. Leprosy can be tuberculoid, which is a localized form exhibiting few or no bacteria, or lepromatous, which is a systemic form with high loads of mycobacteria. Among the tuberculoid patients, there is a range of skin granulomatous diseases phenotypically comparable to leprosy (5).

The use of PCR for leprosy diagnosis has been extensively tested (4,16,33–44). However, limitations towards the experimental designs for some published studies were identified. We observed that most studies: (i) test only samples from leprosy patients, creating difficulties in determining some diagnostic parameters such as specificity; (ii) were performed on small sample sizes; and (iii) do not have independent validation on the same assay or an evaluation of the same protocol in different centers. Furthermore, no studies have used reagents produced under good manufacturing practices (GMP), a set of guidelines that allow for traceability and batch-to-batch reproducibility of characteristics such as physical parameters and performance of the reagents (45).

In this study, we solved some of these issues by (i) developing and validating an assay based on the two most tested targets in the literature with better accuracy so far (7,8,41,46), (ii) following guidelines for validation of diagnostic tests (45,47,48), and (iii) using GMP grade reagents. We were also able to include a reaction for the detection of human 18S gene in the sample, to assess the quality of DNA extraction and reagents performance in the same reaction as the *M. leprae* determination occurs.

RLEP and 16SrRNA are the most frequent markers used in leprosy studies, displaying PCR sensitivity values up to 80% for each target. However, it is important to note that the sensitivity of targets varied between sample types, clinical settings, and also between studies of the same authors (8,9). Tatipally et al. (9) showed that using more than one marker in a multiplex format of conventional endpoint PCR yields significantly higher PCR positivity.

In the currently study, a multiplex qPCR assay simultaneously amplifies two specific *Mycobacterium leprae* targets (16SrRNA and RLEP), and the mammalian 18SrRNA gene as internal reaction control. The assay validation comprised analytical performance, diagnostic sensitivity and specificity, as well as reproducibility and repeatability. Development of multiplex qPCR assays provides a greater challenge than designing singleplex assays because it often requires extensive optimization as primer-dimers and non-specific interactions may interfere with amplification of the desired targets. Additionally, it is important that the amplification of two or more targets does not preferentially amplify one of the targets (49,50). Combining multiple primers and probes did not affect the efficiency of the triplex qPCR in comparison to the corresponding singleplex reactions used in Martinez et al. (16), who evaluated the independent detection of 16S and RLEP using the same primers and probes and obtained 0.91 and 0.51 for sensitivity and 0.73 and 1 for specificity, respectively. Barbieri et al. (4) also used the same 16S target to evaluate paucibacillary leprosy samples and obtained 0.57 for sensitivity and 0.91 for specificity. Here, we evaluated a panel with 53 leprosy and 44 non-leprosy patient samples, and later a different sample panel (50 patient samples) and achieved high sensitivity (> 90%) and specificity (100%) for both panels tested.

However, we understand that the small number of paucibacillary (PB) individuals in our study is a limitation. In fact, the greatest importance of using qPCR as a complementary diagnosis is precisely for PB samples. Generally, PB patients exhibit low (or zero) bacterial load and a histopathology examination that does not distinguish from the diagnosis of other dermatoses. Therefore, these are the cases where clinical evaluation alone might not be able to determine the diagnosis, and where a qPCR confirmation becomes more important. However, due to the scarcity of bacterial DNA in these sample, it is known that the detection of *M. leprae* in PB patients by real-time PCR is difficult (4).

The reactions we developed in this study predict the equivocal classification of early-stage infections based on the finding from Martinez et al. (16), who showed that RLEP displays higher sensitivity than 16S whereas the ribosomal gene displays higher specificity. Thus, samples lacking 16S amplification but with RLEP amplification with a C_p_ lower than the threshold are suggested to be re-analyzed.

In general, our data (figure 6) show a correlation between BI and C_p_ values. Biopsies from patients with higher BI values were deemed positive for bacteria earlier in the amplification cycle, as seen by the lower Cp values and high copy numbers of bacilli.

The “analytical sensitivity” or “limit of detection” of an assay is defined as the ability of the assay to detect very low concentrations of a given substance in a biological specimen (45). The result of the limit of detection (LoD_95%_) determination when tested on a purified *M. leprae* sample indicated a higher sensitivity for RLEP (4.6 fg of DNA/reaction, equivalent to approximately 1.3 *M. leprae* genomes) versus 16S (450 fg of DNA/reaction, approximately 126 *M. leprae* genomes). This difference in sensitivity was expected since the 16SrRNA is a single copy gene (29) and the RLEP presents an average of 36 copies per genome (26).

Applicability in a reference laboratory setting was also considered during the development of these reactions. Intra and inter-operator variability were low, ensuring consistent results in routine testing (table 3). Moreover, reagents remained stable for at least five months, allowing for adequate stock maintenance (figure 4).

Leprosy is a silent disease with a very long incubation time. Currently, transmission can only be halted if patients obtain early diagnosis. High-risk individuals, which are the patients’ close contacts, should be traced and treated whenever leprosy is detected. Recently, it has been suggested that novel policies towards this group of contacts such as immuno- and chemoprophylaxis are effective to help control the disease burden (15,51,52). These approaches provide a screening of the high-risk population that, coupled with a pharmacological or immunological intervention, has been suggested to decrease disease incidence.

In some situations, clinical diagnosis needs the accuracy support of a laboratory analysis, and qPCR is a reliable technique to enable diagnostic confirmation (10). Indeed, we confirmed that the availability of molecular tests can be very helpful in diagnosing patients during contact monitoring (53). When contacts present a leprosy-like lesion, a positive PCR resulted in a leprosy diagnosis with 50% sensitivity and 94% specificity (53). Other indirect methods based on simultaneous detection of host humoral as well as cellular immune response directed against the bacteria are also promising new diagnostic tools. Recently, lateral flow assays (LFA), combining detection of mycobacterial components and host proteins, proved to be specific and sensitive (54–60). The signature detected by this platform identified 86% of the leprosy patients, with a specificity of 90% (AUC: 0.93, p < 0.0001) (58). Thus, a multicentric study comparing different available methods such as qPCR and LFA is still necessary. It is noteworthy that our data showed accuracy, sensitivity, and specificity values quite similar to LFA.

We believe that the diagnosis of tropical and neglected diseases needs molecular-based methods such as PCR, especially due to the robustness and capillarity of the technique in clinical analysis laboratories worldwide. Towards that future, we present a real-time quantitative PCR produced with GMP reagents that adheres to all quality control specifications, allowing batch-to-batch performance reproducibility and repeatability, and that can be used in research and clinical laboratories with reasonable infrastructure in endemic countries. Finally, we envision the multiplex qPCR assay developed adapted to more affordable, rapid, point-of-care tests to be used in low-resourced settings, enabling on-site early and specific diagnosis of leprosy, hopefully helping disease control.

## Data Availability

All data is contained in the manuscript.

## Acknowledgments

The authors are grateful to the entire team of dermatologists, nurses, and technicians that collaborate at the Souza Araújo Clinic from the Leprosy Laboratory at the Oswaldo Cruz Institute. The authors are also grateful for the excellent technical assistance by Aline Burda Farias, Nilson José Fidêncio and Sylvia Mara Bohn at IBMP. We thank the Laboratório de Pesquisa em Leishmaniose (IOC-Fiocruz) and Laboratório de Biologia Molecular Aplicada a Micobactérias (IOC-Fiocruz) for donating the DNA from mycobacterial samples used in the analytical specificity study.

---

**Supplementary Appendix 1**. Sequences of the synthetic DNA template control.

**Supplementary Table 1**. Validation multiplex real-time qPCR assay study results.

**Supplementary Table 2**. Reproducibility and Repeatability results from synthetic DNA.

**Supplementary Table 3**. List of C_p_ cutoff value combinations with associated sensitivity and specificity scores.

**Supplementary Table 4**. Individual Ct values for targets included in the multiplex real-time qPCR assay (16S rRNA/RLEP/18SrRNA), sociodemographic, and laboratory variables for patients samples included in this study.

## Author’s contributions

F.S.N.M., T.J. Wrote the manuscript and contributed equally for this study;

F.S.N.M., T.J., R.C.P.R., N.Z., S.M., M.R.A. designed, performed, and analyzed experiments;

F.S.N.M., M.O.M. supervised sample collection;

T.J., A.D.T.C. supervised production of kit prototypes;

M.A.K., M.O.M. secured funding;

A.D.T.C., M.O.M. conceptualized the study and approved the final version of the manuscript and are co-senior authors of this study. Guarantors of the integrity of the data presented.

